# Using risk prediction models to inform personalized, cost-effective treatment recommendations

**DOI:** 10.1101/2025.08.07.25333118

**Authors:** Mariana R. Neves, Molly Franke, Carole Mitnick, Nelly Ciobanu, Valeriu Crudu, Jennifer Furin, Ted Cohen, Reza Yaesoubi

## Abstract

**Objective:** For many medical conditions, rapid, reliable, and affordable diagnostic tests are not available, which leads clinicians to base treatment decisions on patient symptoms and history. Although prediction models can estimate disease risk, they typically do not account for the downstream health or cost consequences of acting on their predictions. We developed and evaluated methods that integrate risk prediction with decision modeling to inform personalized, cost-effective treatment recommendations.

**Materials and Methods:** We considered two integration methods to maximize the population net monetary benefit (NMB), which summarizes both health and cost outcomes of available actions. In the *probability-based* method, the predicted probability of disease from a risk prediction model is used as input to a decision model. In the *classification-based* method, the decision model relies on the risk prediction model’s binary disease classification. We applied these methods to select between two treatment regimens for patients with rifampicin-resistant tuberculosis in Moldova, while accounting for cost, toxicity, and efficacy associated with each regimen.

**Results:** Both integration methods yielded higher population NMB than standard of care and approaches based on fixed classification thresholds (e.g., 50% or the threshold that maximizes the Youden’s index). However, the classification-based approach was less sensitive to whether the model predictions were properly calibrated.

**Conclusion:** Integrating risk predictions with decision models offers a principled framework for making personalized, value-based treatment decisions. These methods explicitly account for health and cost consequences of treatment choices informed by risk prediction models, improving care quality and resource use in settings with diagnostic uncertainty.

## Introduction

Rapid, reliable, and affordable diagnostic tests are not always available to detect various medical conditions. Consequently, clinicians often rely on the characteristics and symptoms of patients to guide treatment decisions. For example, waiting for a definitive diagnosis of certain infectious diseases (including urinary tract infections, gonorrhea, and chlamydia) could delay therapy; hence, initial treatment of these conditions is empiric, lacking information on the causative agent [1, 2]. This results in many patients receiving ineffective treatment or being unnecessarily treated.

Risk prediction models can be used to predict the presence of a disease (or the causative agent of the disease) based on the characteristics of the patient observable at the time of diagnosis, such as age, symptoms, and other risk factors [3]. While potentially accurate and useful for diagnosis, risk prediction models do not account for the effectiveness, side effects, and long-term health and cost outcomes of actions (e.g. treat versus not treat) that could be informed by the risk prediction. Hence, risk prediction models alone are not sufficient to inform clinical decisions.

In this study, we describe methods to integrate risk prediction models that predict the presence of a disease based on patient characteristics, with decision models that account for long-term health and the cost consequences of available treatment options. For a given patient, these risk-informed decision models aim to identify the treatment option with the highest expected net monetary benefit (NMB), which summarizes both health and cost consequences into a single measure. We also describe how to account for two critical sources of uncertainty when informing decisions: 1) the uncertainty in the performance measures of the risk prediction model (e.g., sensitivity and specificity) due to limited size of the training dataset, and 2) the uncertainty in parameter estimates used in the decision model (e.g., cost, side effects, and effectiveness of treatment options) to project the health and cost consequences of available options.

We apply the proposed methods to select between two treatment regimens for patients with rifampicin-resistant tuberculosis (RR-TB) in Moldova. We assess whether the personalized treatment regimens recommended by the proposed methods are expected to increase the overall NMB of patients with RR-TB compared to the status quo.

## Methods

### Problem formulation

We consider a scenario where each member of a population could have a disease. The presence of the disease is not known with certainty (e.g., due to the lack of accurate diagnostic tools). Effective treatments are available, but can be costly and/or can cause side effects. If left untreated, the disease can cause adverse health and/or financial outcomes. In this scenario, clinicians often rely on the patient’s symptoms and other risk factors to decide whether to treat the patient. The choice between treatment options can be formulated using the decision model presented in Fig. 1, where treatment options *A*_1_ and *A*_2_ are assumed to be available. We note that in this context, *A*_1_ and *A*_2_ may represent “no treatment” versus “treatment”, although they could alternatively represent two different treatment types, depending on the clinical setting.

**Figure 1:**
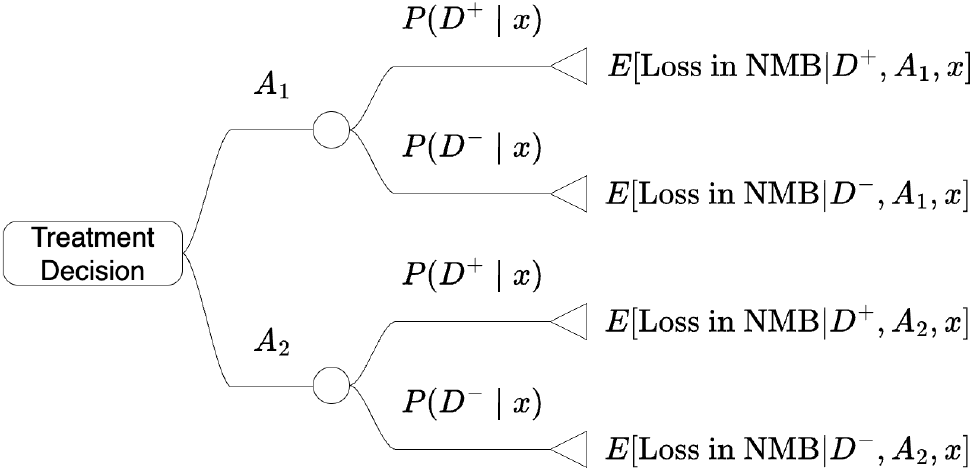
A decision tree to decide between treatment options *A*_1_ and *A*_2_ for an individual with observable characteristics *x* who may or may not have a disease.

The health and financial outcomes of treatment options depend on various factors such as the effectiveness, side effects, and cost of treatment options. The effectiveness of a treatment option depends on the true disease state *D* ∈ {*D*^+^, *D*^−^}, where *D*^+^ represents the presence of the disease and *D*^−^ represents the absence of the disease. For an individual with observable characteristics *x*, we use E[*C* | *D, A, x*] and E[*Q* | *D, A, x*] to denote the expected cost and expected health loss if the patient’s true state of disease is *D* ∈ {*D*^+^, *D*^−^} and the treatment option *A* ∈ {*A*_1_, *A*_2_} is selected. The expected cost and health loss under each disease and treatment option pair (*D, A*) (i.e., E[*C* | *D, A, x*] and E[*Q* | *D, A, x*]) is influenced by the effectiveness, cost, side effects of the treatment and the adverse health and cost outcomes associated with the disease.

To evaluate each treatment option, we use the loss in net monetary benefit (NMB), which incorporates both the expected cost and expected health loss into a composite measure [4]. For a patient with observable characteristics *x*, the expected loss in NMB for a disease and treatment option pair (*D, A*) is defined as:

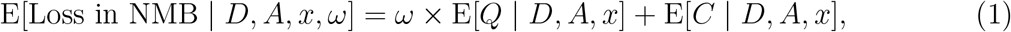

where *ω* is the decision-maker’s willingness-to-pay (WTP) for one unit of health (e.g., per disability-adjusted life-years averted).

To select the optimal treatment, we calculate the expected loss in NMB under each treatment option *A* ∈ {*A*_1_, *A*_2_} by marginalizing over the disease state *D* ∈ {*D*^+^, *D*^−^}; that is

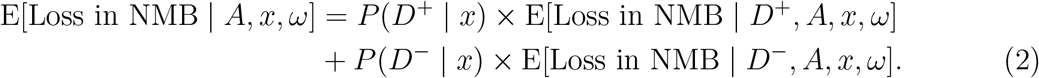

In Eq. (2), *P* (*D*^+^ |*x*) is the probability that the patient has the disease and *P* (*D*^−^ *x*) = 1 − *P* (*D*^+^| *x*) is the probability that the patient is disease-free. For a patient with observable characteristics *x*, the optimal treatment *A*^*^(*x, ω*) is identified by:

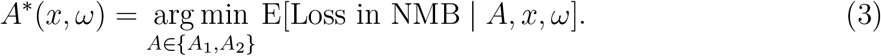

When the probability *P* (*D*^+^ | *x*) is not available for each individual, the probability of the disease being present is assumed to be independent of the observable characteristics of the patient and the estimated prevalence of the disease among the population of interest is often used as a proxy. If an individual-level risk prediction model is available, the outcome of this model could be integrated into the decision model in Fig. 1 to inform personalized and cost-effective treatment recommendations. We refer to decision models that are informed by risk prediction models as Prediction-Informed Decision Models (PIDEMs).

### Prediction-Informed Decision Models (PIDEMs)

Suppose that a risk prediction model that estimates the probability of disease for a patient with observable characteristics *x* is available. These characteristics may include demographic factors (e.g., age, sex), clinical information (e.g., symptom type, medical history), and contextual variables (e.g., geographic location). We use 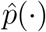 to represent this risk prediction model, which for a patient with characteristics *x* estimates the probability of disease as 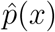. We consider two methods to use 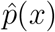 to inform the decision model in Fig. 1, which differ in how 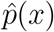 is integrated into the model. For the first method, which we refer to as *probability-based* PIDEM (PIDEMp), we use 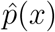 directly as an estimate for *P* (*D*^+^ | *x*) in Fig. 1.

For the second method, which we refer to as *classification-based* PIDEM (PIDEMc), we compared 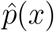 with a classification threshold *t* ∈ [0, 1]; if 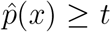, the patient is classified as having the disease, which we denote by *T* ^+^; otherwise, the patient is classified as disease-free, which we denote by *T* ^−^. To decide whether to treat or not to treat a patient based on the classification results, we replicate the decision model in Fig. 1 to create two decision models (Fig. 2): one for when the individual is classified as having the disease (left panel in Fig. 2) and one for when the individual is classified as disease-free (right panel in Fig. 2). These decision models are identical except for the probability of disease. In the left-hand side decision model, the disease probability is estimated by *P* (*D*^+^ | *T* ^+^; *t*), which represents the probability of disease if the patient is classified as *T* ^+^ at classification threshold *t*; and in the right-hand side decision model, the disease probability is estimated by *P* (*D*^+^ | *T* ^−^; *t*), which represents the probability of disease if the patient is classified as *T* ^−^.

**Figure 2:**
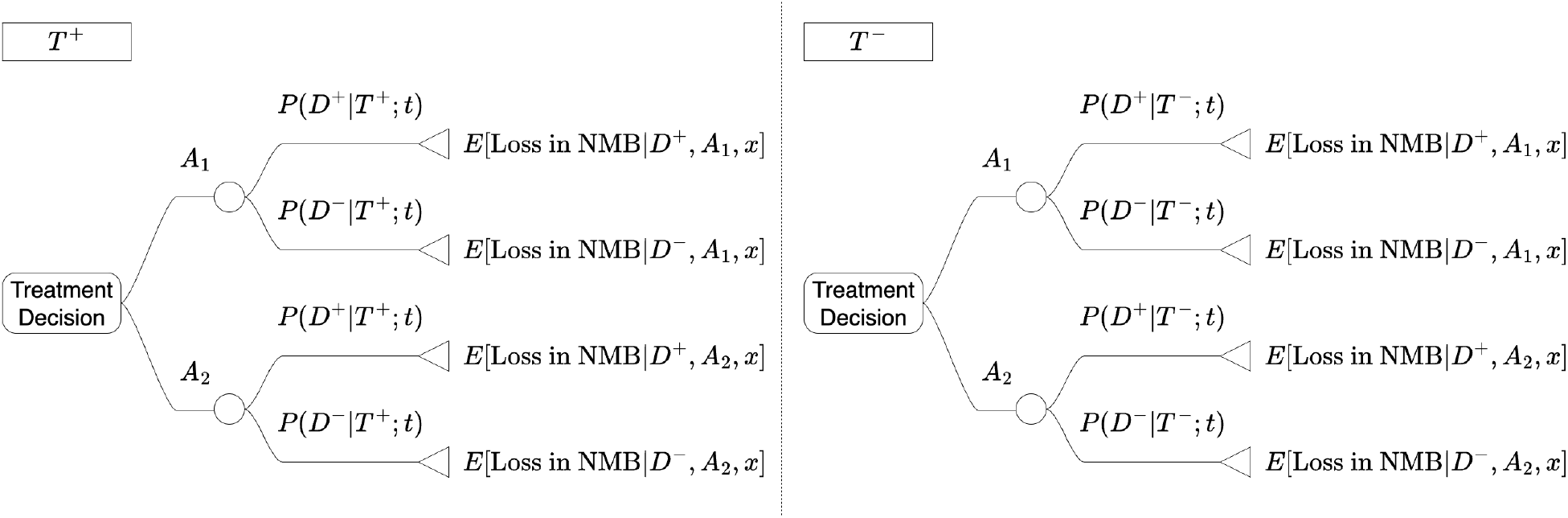
Decision trees used for selecting the optimal treatment for an individual classified as having the disease (denoted by *T* ^+^, left panel) and for an individual classified as disease-free (denoted by *T* ^−^, right panel). Variable *t* is the classification threshold used by the risk prediction model.

The probabilities *P* (*D* | *T*; *t*), *D* ∈ {*D*^+^, *D*^−^}, *T* ∈ {*T* ^+^, *T* ^−^} can be estimated using the Bayes’ theorem and the sensitivity and specificity of the risk prediction model at each threshold *t* ∈ [0, 1], which we denote by *Se*(*t*) and *Sp*(*t*). The probability that the disease is present given classification results *T* ^+^ can be calculated as:

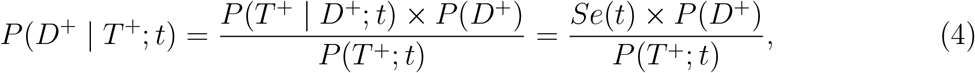

and the probability that the disease is absent given classification results *T* ^−^ can be calculated as:

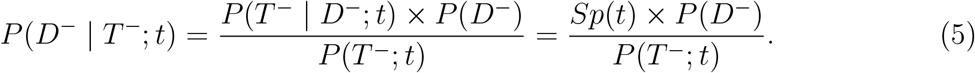

In the decision trees in Fig. 2, we can set *P* (*D*^−^ | *T* ^+^; *t*) = 1 − *P* (*D*^+^ | *T* ^+^; *t*) where *P* (*D*^+^ | *T* ^+^; *t*) is calculated in Eq. (4) and set *P* (*D*^+^ | *T* ^−^; *t*) = 1 − *P* (*D*^−^ | *T* ^−^; *t*) where *P* (*D*^−^ | *T* ^−^; *t*) is calculated in Eq. (5).

### Incorporating the uncertainties in the predicted probability of disease

To identify the optimal treatment option, the PIDEMp method uses 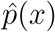 as an estimate of the probability of disease (i.e, *P* (*D*^+^ | *x*)). For a patient with observable characteristics *x*, the optimal treatment choice is the one with the lowest expected loss in NMB calculated in Eq. (2), which depends on *P* (*D*^+^ | *x*) and is not affected by the uncertainty in 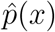.

In contrast, the PIDEMc method uses *P* (*D*^+^ |*T* ^+^; *t*) and *P* (*D*^+^ |*T* ^−^; *t*) as estimates of the probability of disease given the classification result *T* ^+^ or *T* ^−^. These probabilities depend on the sensitivity and specificity of the risk prediction model (through Eq. (4) and Eq. (5)), which are subject to uncertainty and are affected by, for example, the size of the dataset and the distribution of positive and negative outcomes in the dataset. The uncertainty in sensitivity and specificity could propagate affecting *P* (*D*^+^|*T* ^+^; *t*) and *P* (*D*^+^|*T* ^−^; *t*).

To mitigate this issue and incorporate uncertainties in estimates for *P* (*D*^+^ |*T* ^+^; *t*) and *P* (*D*^+^ |*T* ^−^; *t*), we use a bootstrapping approach that is commonly used to correct for optimism in estimates for performance metrics, such as area under the receiver operating characteristic (auROC) curve, sensitivity, and specificity [5].

Let *X* be the entire dataset and 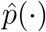, as previously defined, be the risk prediction model trained on *X* using feature selection, and hyperparameter tuning methods to minimize over-fitting. Let *µ* be the performance metric we aim to estimate for the risk prediction model 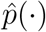, such as the auROC curve. An estimate of *µ* can be obtained by evaluating the risk prediction model 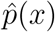 against the dataset *X*. This estimate is commonly referred to as “apparent” performance, and we denote it by 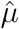. Since 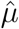 is calculated using the same dataset that is used to develop the model, it could be optimistic in estimating the risk prediction model’s performance.

To characterize the distribution of optimism in 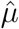, we obtain *M* bootstrap samples, with replacement, from *X* to create bootstrap datasets *X*_1_, *X*_2_, …, *X*_*M*_ (all of the same size as For each bootstrap dataset *X*_*i*_, we then develop a new risk prediction model 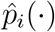 using appropriate feature selection and hyperparameter tuning methods. Let 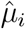 be the apparent performance of this model, which is calculated with the bootstrap dataset *X*_*i*_ and 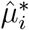 be the performance of this model calculated with the original dataset *X*. The optimism of this bootstrap dataset is 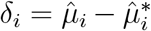. We can then use the sample 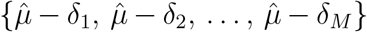 to estimate the risk prediction model’s performance *µ*, with the optimism-corrected estimate given by 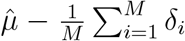. The confidence interval can be computed as 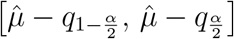, where *q*_*p*_ denotes the *p* × 100-th percentile of the sample *δ*_1_, …, *δ*_*M*_. The pseudocode for this procedure is presented in Algorithm 1.

An issue that may arise when using this approach to estimate *P* (*D*^+^ |*T* ^+^; *t*) or *P* (*D*^+^ |*T* ^−^; *t*) is that estimates of these probabilities may not be defined for all bootstrap datasets. This occurs when some bootstrap risk prediction model 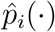 results in classifying all patients in the dataset *X*_*i*_ as *T* ^+^ or *T* ^−^ for the selected classification threshold *t*. To address this issue, we note that as *t* approaches 1 fewer patients are classified as *T* ^+^ and the chance that *P* (*D*^+^ | *T* ^+^; *t*) is not defined increases. Therefore, if for some *t, P* (*D*^+^ | *T* ^+^; *t*) cannot be estimated, we use the estimate of *P* (*D*^+^ | *T* ^+^; *t*^*′*^) for the largest *t*^*′*^ *< t* that was obtained with at least 20 individuals classified as *T* ^+^. Likewise, we note that as *t* approaches 0, the chance that *P* (*D*^−^ | *T* ^−^; *t*) is not defined increases since fewer patients are classified as *T* ^−^. Therefore, if for some *t, P* (*D*^−^ | *T* ^−^; *t*) cannot be estimated, we use the estimate of *P* (*D*^−^ | *T* ^−^; *t*^*′*^) for the smallest *t*^*′*^ *> t* that was obtained with at least 20 individuals classified as *T* ^−^. Finally, we note that Algorithm 1 calculates three components (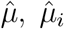 and 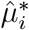) to generate optimism-corrected estimates (Step 10). If any of these components cannot be calculated, we use this approach to approximate that component for the given classification threshold *t*.

### Incorporating the uncertainties in cost and health-related parameters of the decision model

Using the decision models in Fig. 1 and Fig. 2 to inform decisions for a patient with observable characteristics *x* requires estimating the expected loss in NMB for all disease and treatment option pairs (*D, A*) (i.e., E[Loss in NMB | *D, A, x, ω*] with *D* ∈ {*D*^+^, *D*^−^} and *A* ∈ {*A*_1_, *A*_2_} as defined in Eq. (1)). For this purpose, simulation models are often used to project the loss in NMB under different treatment options [6]. These projections are subject to uncertainty due to the inherent uncertainties in model parameters (e.g., cost items and quality of life in different disease states) that influence the future cost and health outcomes associated with alternatives under consideration.

#### Algorithm 1

Correcting for optimism in estimates for performance measures of the risk prediction model (e.g., auROC).

**Requires**: Entire dataset *X*.

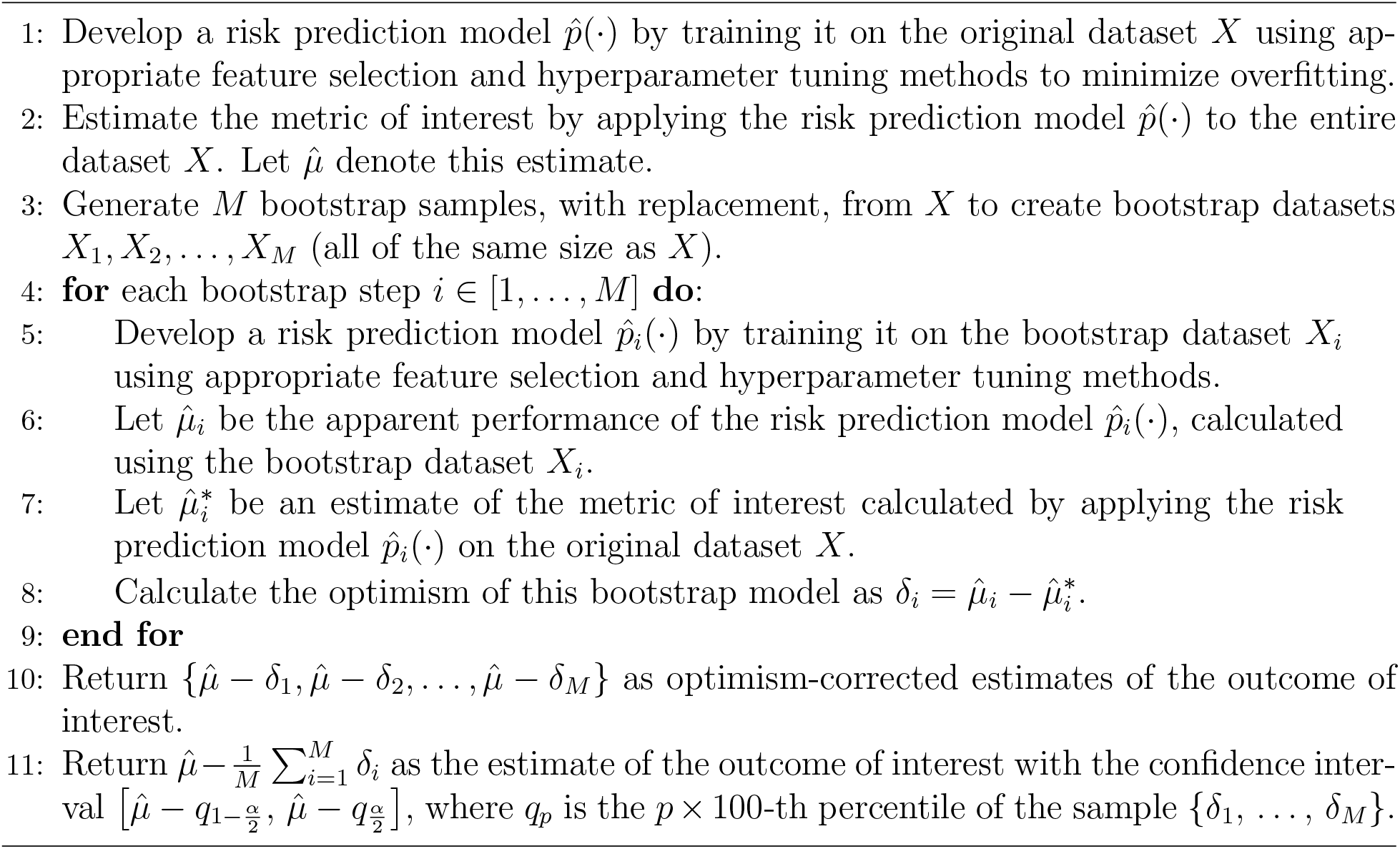

The common approach to propagating uncertainties from model parameters to the estimated costs and effect of treatment alternatives is probabilistic analysis, also known as probabilistic sensitivity analysis [7, 8]. In this approach, we assign appropriate probability distributions to uncertain model parameters such as parameters related to cost, quality of life, and duration of various health states. We then draw many random samples (*S >* 1) from these parameter distributions, denoted by {*θ*_1_, *θ*_2_, …, *θ*_*S*_} and use the simulation model to obtain estimates of E[Loss in NMB|*D, A, x, ω*] under each sampled parameter *θ*_*i*_. We denote these samples by *l*_*i*_(*D, A, x, ω, θ*_*i*_). In the next section, we describe how these estimates are incorporated to inform treatment decisions for a given patient.

### Using a PIDEMp to inform decisions

Suppose that the parameter samples {*θ*_1_, *θ*_2_, …, *θ*_*S*_} are obtained through the probabilistic sensitivity analysis described in the previous section and a WTP threshold *ω* is set by the decision-maker. Under PIDEMp, we estimate *P* (*D*^+^ | *x*) in Fig. 1 with 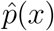. Using each parameter sample *θ*_*i*_, the expected loss in NMB under each option *A* ∈ {*A*_1_, *A*_2_} can be obtained by marginalizing *l*_*i*_(*D, A, x, ω, θ*_*i*_) over the disease state *D* (as in Eq. (2)):

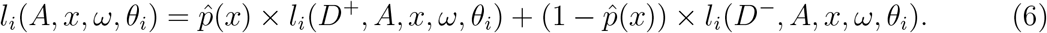

We then estimate E[Loss in NMB|*A, x, ω*] with 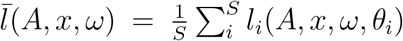. This allows us to find the optimal treatment option using Eq. (3). Algorithm 2 provides the pseudo-code for this approach.

#### Algorithm 2

Using PIDEMp to find the optimal treatment option and the associated expected loss in NMB for a patient with observable characteristics *x*.

**Requires**: Risk prediction model 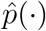, WTP value *ω*, and parameter samples {*θ*_1_, …, *θ*_*S*_}.

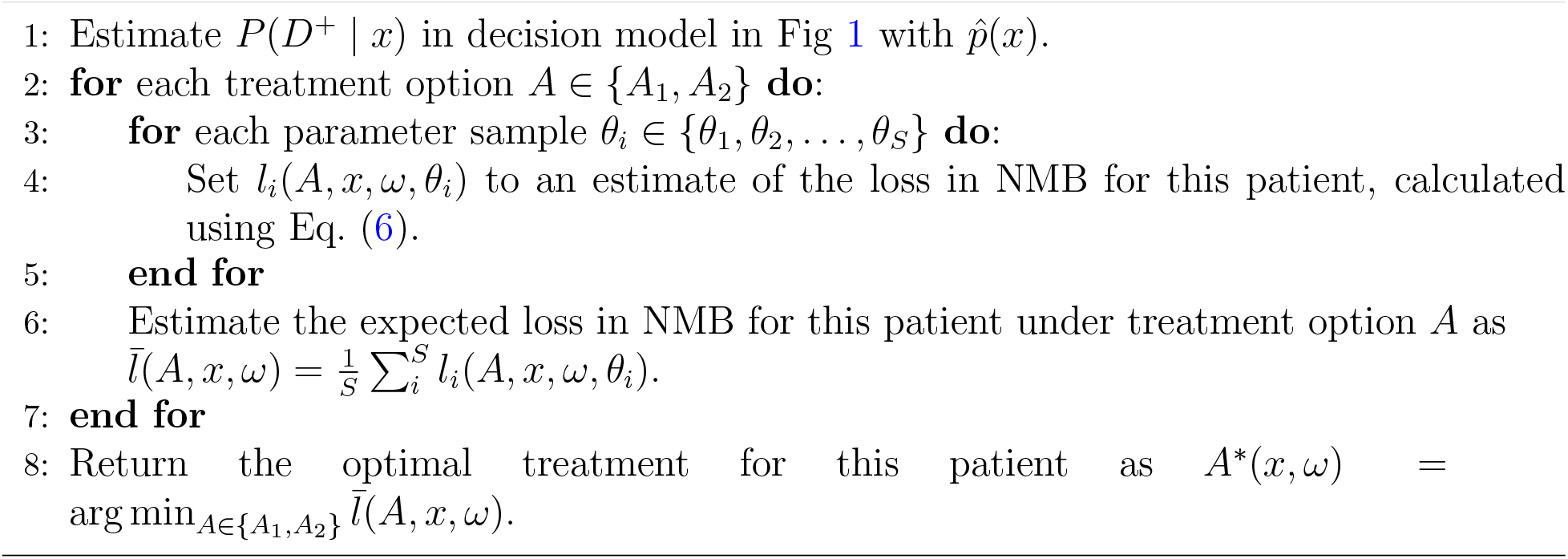

### Using a PIDEMc to inform decisions

In the PIDEMc approach, we use the risk prediction model 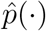 to classify a patient as *T* ^+^ (if 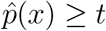) or *T* ^−^ (if 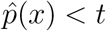). We again assume that the parameter samples {*θ*_1_, …, *θ*_*S*_} are obtained through the probabilistic sensitivity analysis and the WTP threshold *ω* is set by the decision-maker. To incorporate uncertainties in the predicted probability of disease, we apply Algorithm 1 to create samples 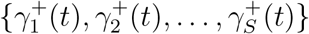 of *P* (*D*^+^|*T* ^+^(*t*)), and samples 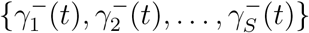 of *P* (*D*^−^|*T* ^−^(*t*)).

If the patient is classified as *T* ^+^, we use the decision model *T* ^+^ in Fig. 2 to inform decisions. To this end, we use each model parameter sample *θ*_*i*_ and probability sample 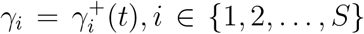 to estimate the expected loss in NMB under each treatment option *A* ∈ {*A*_1_, *A*_2_} (as in Eq. 2):

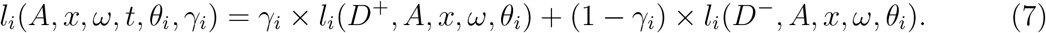

We then estimate the expected loss in NMB under the treatment option *A* as 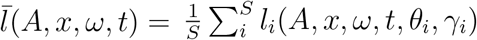. We can use Eq. (3) to find the optimal treatment option.

If the patient is classified as *T* ^−^, we use the decision model *T* ^−^ in Fig. 2 to select the optimal treatment option. In this case, we calculate the expected loss in NMB under each treatment option *A* ∈ {*A*_1_, *A*_2_} using the Eq. (7) but with 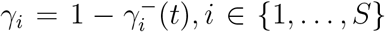. Algorithm 3 provides the pseudo-code for this approach.

#### Algorithm 3

Using PIDEMc to find the optimal treatment and the associated expected loss in NMB for a patient with observable characteristics *x*.

**Requires**: Risk prediction model 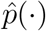, WTP value *ω*, classification threshold *t*, samples {*θ*_1_, …, *θ*_*S*_} of the parameters of the decision model to estimate losses in NMB, samples 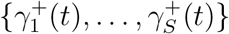 of the probability *P* (*D*^+^|*T* ^+^(*t*)), and samples 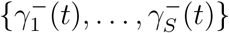 of the probability *P* (*D*^−^|*T* ^−^;*t*) (obtained using Algorithm 1)

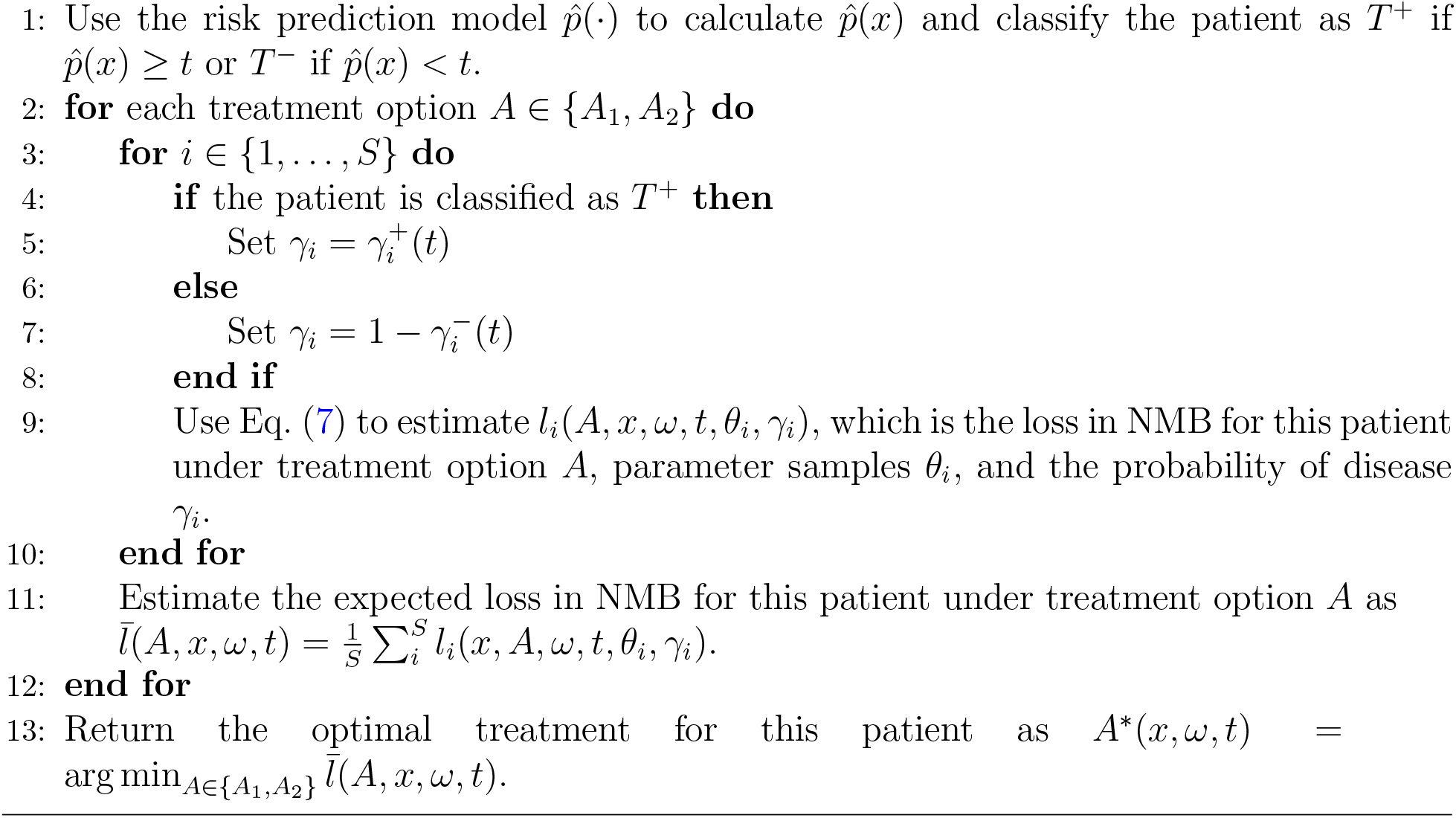

### Optimizing the classification threshold *t*

The algorithm for using PIDEMc to inform decisions (Algorithm 3) assumes that a classification threshold *t* is provided. The choice of *t* determines the sensitivity and specificity of a risk prediction model. Sometimes, *t* is arbitrarily selected as 0.5. However, if false negatives and false positives have different levels of importance for the decision maker, the threshold *t* could be selected so that it minimizes the most critical error, potentially with constraints on the other. Another approach is to choose the threshold *t* to maximize the Youden’s index, which is defined as *Se*(*t*) + *Sp*(*t*) − 1 [9]. Nonetheless, none of these approaches to select the classification threshold *t* account for the costs and health impact of false negatives and positives.

We propose to determine the classification threshold *t* such that the overall loss in NMB among the patients included in our dataset is minimized. To find the optimal classification threshold, we first select a large number of thresholds between 0 and 1: {*t*_1_, *t*_2_, …, *t*_*T*_}. Then, for each threshold *t*∈ {*t*_1_, *t*_2_, …, *t*_*T*_}, we estimate the expected loss in the average NMB per patient if Algorithm 3 is used at this threshold to inform decisions for all patients in our dataset *X*. Algorithm 4 details the steps for selecting the optimal threshold *t*^*^.

#### Algorithm 4

Optimizing the classification threshold *t* to minimize the loss in the population NMB.

**Requires**: Entire dataset *X*, risk prediction model 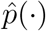, WTP value *ω*, samples {*θ*_1_, *θ*_2_, …, *θ*_*S*_} of parameters of the simulation model to estimate losses in NMB, samples 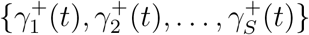 of the probability *P* (*D*^+^ | *T* ^+^(*t*)), and samples 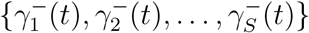 of the probability *P* (*D*^−^ | *T* ^−^(*t*)) (obtained from Algorithm 1).

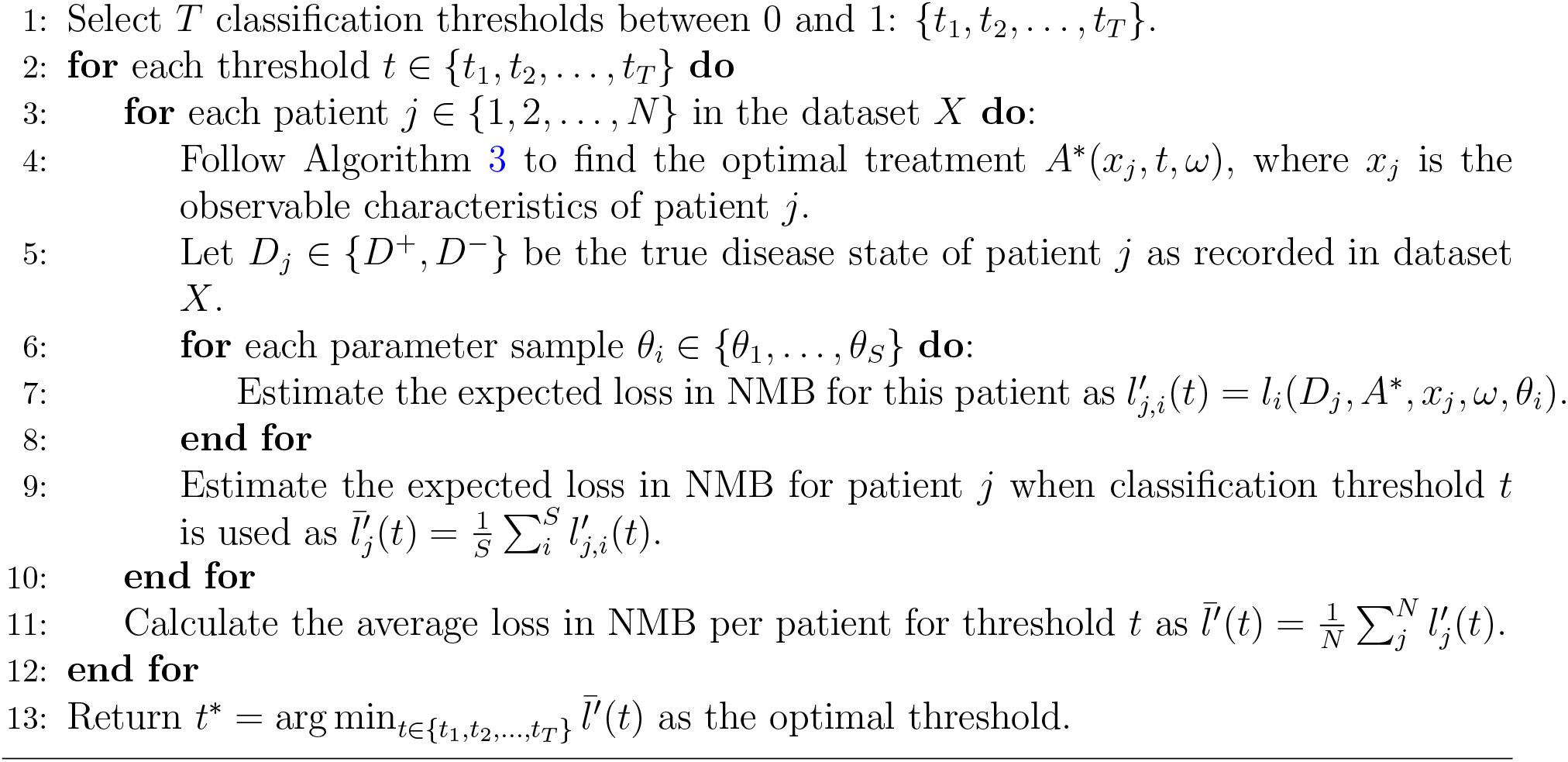

### Case Study: Identifying Personalized and Cost-Effective Treatment Regimens for Patients with Rifampicin-Resistant Tuberculosis

We applied the methodology described in this paper to identify cost-effective treatment regimens for patients with rifampicin-resistant tuberculosis (RR-TB) based on their basic demographic and clinical information, using an illustrative example. While the rate of TB cases has declined worldwide [10], current efforts to control TB epidemics are challenged by the emergence of *M. tuberculosis* strains that are resistant to essential antibiotics in the standard TB treatment regimen [11]. With the widespread use of Xpert MTB/RIF (a molecular test for the rapid detection of TB and resistance to rifampicin) over the last 10 years, a growing number of individuals with RR-TB are being detected in many high-burden TB settings [12]. Due to limited access to drug-susceptibility tests (DSTs) and long delays in receiving DST results (up to 12 weeks) [13, 14], the treatment of individuals with RR-TB remains empiric (i.e., without the results of DSTs) and according to standardized regimens, which are determined at the global level [2].

In 2022, the World Health Organization (WHO) endorsed a novel treatment regimen for RR-TB, known as BPaLM [2], composed of bedaquiline, pretomanid, linezolid and moxifloxacin, the latter belonging to the fluoroquinolone (FQ) class of antibiotics. However, recent rises in the prevalence of resistance to FQs [15, 16] have challenged the effectiveness of this regimen. In this context, a key question is whether certain patients with RR-TB should be treated with a modified version of the BPaLM regimen in which FQ is replaced with clofazimine (CFZ), which is another important second-line antibiotic with a lower prevalence of resistance but more expensive. We refer to this modified BPaLM regimen as BPaLC. Although not yet formally recommended by the WHO, the BPaLC regimen has been evaluated in clinical trials [17], and robust data are available to support its use in our illustrative example.

We applied the method described in this paper to decide between BPaLM and BPalC given specific patient characteristics (such as age, residence in urban or rural areas, and their history of TB treatment). We estimate the average loss in NMB per patient with RR-TB under the status quo, where all patients with RR-TB are prescribed BPaLM, and under the scenario where PIDEMp and PIDEMc are used to decide between BPaLM and BPaLC given the patient’s characteristics. Although the numbers used (costs, DALYs, and probabilities) correspond to RR-TB treatments, this example is illustrative and has been simplified to demonstrate the methodology proposed in this paper.

### Data source and study population

To develop a risk prediction model that predicts resistance to FQs, we used a dataset from the Republic of Moldova comprising 540 patients diagnosed with TB notified between January 2018 and December 2019, featuring 32 variables containing demographic and clinical information. The dataset includes only patients with RR-TB who had at least one conclusive test result, either positive or negative, for additional resistance to FQs. A detailed description of the dataset is provided in [18].

### A Logistic Regression Model to Predict Resistance to FQ

To estimate the probability that a RR-TB demonstrates resistance to FQ, we use logistic regression models with *l*_2_ regularization [19] and 10-fold cross validation for hyperparameter tuning with the goal of maximizing the auROC curve. The model incorporates various demographic factors, including age, sex, family size, occupation, education, living condition, residency history outside Moldova, urban living status, homelessness, information on financial assistance, prior incarceration history and district prevalence of TB resistant to FQ. Furthermore, we integrated information on the type of TB (e.g., new case versus relapse) and the location of infection (e.g., pulmonary vs. extrapulmonary) into the model. The definition of these variables and the distribution of values these variables take are presented in [18].

We used the Platt scaling [20] and the beta calibration [21, 22] methods to calibrate the model predictions. We compare these methods using calibration plots, Brier score and the expected calibration error to decide which calibration method to use in the case study analyses.

### PIDEMp and PIDEMc to Inform Treatment Regimens for Patients with RR-TB

To use PIDEMp and PIDEMc to inform treatment regimens for a patient with RR-TB, we let *D*^+^ and *D*^−^ denote resistance and susceptibility to FQs, and *A*_2_ and *A*_1_ denote treatment with BPaLC and BPaLM, respectively. We also need to estimate E[Loss in NMB|*A, D, x*] for a patient with observable characteristics *x* who receives treatment *A* ∈ {*A*_1_, *A*_2_} in disease state *D*∈ {*D*^+^, *D*^−^}. For this illustrative example, we use a simple probability tree (Fig. 3) to simulate the outcomes of each regimen for each disease state. In this probability tree, outcomes of treatment could be 1) Treatment Success, 2) Treatment Failure, and 3) Death. When the patient completes the treatment successfully, there is still a chance of recurrent TB. For this reason, we include two possible outcomes after treatment success: cure and recurrence. The probabilities of these outcomes under different resistance statuses and treatment regimens are provided in Table 1.

**Table 1:** Probabilities of treatment outcomes under different combinations of disease state and treatment regimens (see Fig. 3)

| Disease State<sup>1</sup> | Treatment Regimen | Primary Treatment Outcome (PTO) | PTO Probability<sup>2</sup> | Secondary Treatment Outcome (STO) | STO Probability<sup>2</sup> | Reference |
| --- | --- | --- | --- | --- | --- | --- |
| FQ-R ($D^+$) | BPaLC ($A_2$) | Treatment Success<br>Failure<br>Death | 80%<br>19%<br>1% | Cure<br>Recurrence | 95%<br>5% | [32]<sup>3</sup> |
| FQ-R ($D^+$) | BPaLM ($A_1$) | Treatment Success<br>Failure<br>Death | 75%<br>25%<br>5% | Cure<br>Recurrence | 95%<br>5% | None<sup>4</sup> |
| FQ-S ($D^-$) | BPaLC ($A_2$) | Treatment Success<br>Failure<br>Death | 80%<br>19%<br>1% | Cure<br>Recurrence | 95%<br>5% | [32]<sup>3</sup> |
| FQ-S ($D^-$) | BPaLM ($A_1$) | Treatment Success<br>Failure<br>Death | 89%<br>11%<br>0% | Cure<br>Recurrence | 99%<br>1% | [32]<sup>3</sup> |

<sup>1</sup> FQ-R: resistance to FQs; FQ-S: susceptibility to FQs.
<sup>2</sup> To account for uncertainty in the probability of different outcomes we used a Dirichlet distribution with parameters equal to the samples in each outcome category. When the source study provided only probabilities of each outcome we use 100 times the probabilities as parameters of the Dirichlet distribution.
<sup>3</sup> Probability of outcomes obtained from Table 2 in [32]. We used the estimates for the modified intention-to-treat population at 72 weeks and considered failure a combination of lost to follow-up and early discontinuation of the treatment (due to adherence issues, adverse event, not meeting inclusion or meeting exclusion criteria or withdrew consent during treatment) and treatment failure. Given that BPaLC regimen does not include FQ, we assumed no difference in outcomes for FQ-R and FQ-S under BPaLC.
<sup>4</sup> We assumed that BPaLM would be less effective in treating FQ-resistant TB and, for this patient group, also less effective than BPaLC. Consequently, we assumed higher probabilities of treatment failure and death for BPaLM in cases of FQ-resistant TB. Additionally, we assumed that among successfully treated patients, the probability of recurrence would be the same as that observed with BPaLC.

**Figure 3:**
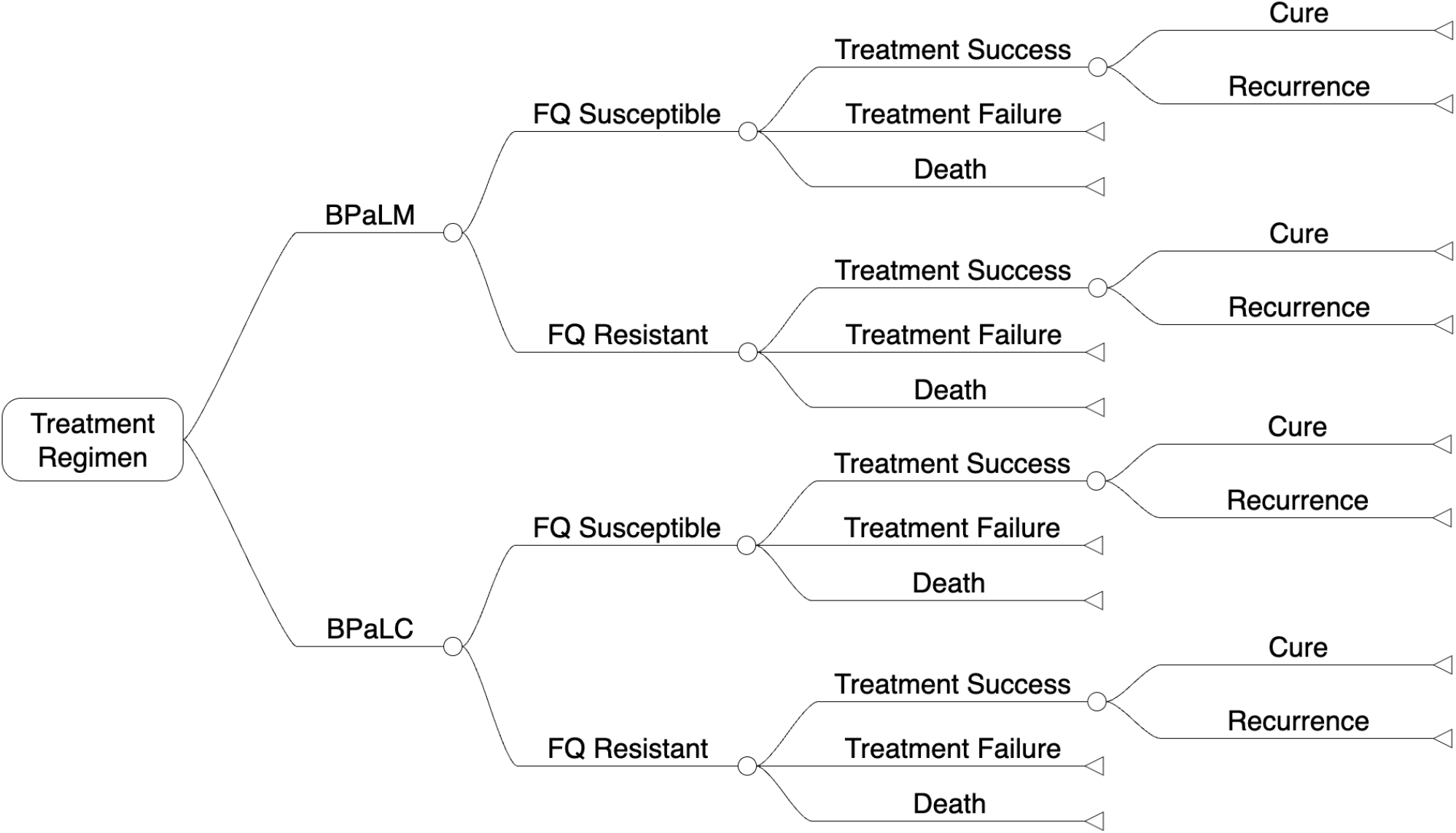
A decision tree representing the treatment and outcomes of patients with RR-TB.

We accounted for costs associated with the BPaLM and BPaLC regimens and unsuccessful treatment outcomes, as described in Table 2. We used gamma distributions to capture the uncertainty in estimates for the cost items (Table 2). We assumed that if the treatment is ineffective or recurrence occurs after treatment (with either BPaLM or BPaLC) the recommended treatment regimen will be replaced by a personalized regimen, which lasts 18 months and costs 10%more than BPaLC. The personlized regimen is determined based on the results of second-line phenotypic DST.

**Table 2:** Costs associated with treatment regimens and unsuccessful treatment outcomes.

| Parameter | Distribution | Reference |
| --- | --- | --- |
| Cost of bedaquiline treatment, first month only | Gamma($\mu = \$126.7$, $\sigma = 0.1\mu$) | [30] |
| Cost of bedaquiline treatment, per subsequent month | Gamma($\mu = \$47.12$, $\sigma = 0.1\mu$) | |
| Cost of pretomanid treatment, per month | Gamma($\mu = \$60.80$, $\sigma = 0.1\mu$) | |
| Cost of linezolid treatment, per month | Gamma($\mu = \$22.44$, $\sigma = 0.1\mu$) | |
| Cost of moxifloxacin treatment, per month | Gamma($\mu = \$4.86$, $\sigma = 0.1\mu$) | |
| Cost of clofazimine treatment, per month | Gamma($\mu = \$19.89$, $\sigma = 0.1\mu$) | |
| Cost per *M. tuberculosis* culture, to be conducted monthly | Gamma($\mu = \$8.04$, $\sigma = 0.1\mu$) | |
| Cost per second-line phenotypic DST, to be conducted if treatment failure or recurrence happens | Gamma($\mu = \$51.29$, $\sigma = 0.1\mu$) | |
| Cost of monitoring side effects, per month<sup>1</sup> | Gamma($\mu = \$69.19$, $\sigma = 0.1\mu$) | [31] |

<sup>1</sup>Obtained from Table S4 in [31]. We estimated the cost of monitoring side effects with a sum of outpatient treatment visit costs (treatment monitoring costs) and laboratory test costs (ECG, Full haemogram, Creatinine Clearance, Lipase and Liver Function Tests) selected to match side effects considered. Costs were obtained in Minsk, Belarus in BYN and were converted to 2021 USD using the conversion rate 1 USD = 2.53 BYN.

We assumed that patients with RR-TB experience a reduced quality of life due to TB symptoms. We use the disability-adjusted life-year (DALY) metric to measure the unit of health loss due to these causes. We assumed TB symptoms are present for the first 8 weeks of the treatment if it is successful. If treatment is unsuccessful or if recurrence occurs, we assumed the patient will continue to experience TB symptoms for the entire course of treatment. The disability weights and the duration of these health outcomes were modeled using probability distributions listed in Table 3.

**Table 3:** Disability weights and the duration of health outcomes.

| Parameter | Distribution | Reference |
| --- | --- | --- |
| Disability weight associated with TB | Beta($\mu = 0.333$, $\sigma = 0.1\mu$) | [33] |
| Years with lower quality of life after |  | [32]<sup>1</sup> |
| Treatment success under BPaLM, no recurrence | Gamma($\mu = 0.167$, $\sigma = 0.1\mu$) | |
| Treatment success under BPaLC, no recurrence | Gamma($\mu = 0.167$, $\sigma = 0.1\mu$) | |
| Treatment success under BPaLM, recurrence | Gamma($\mu = 2.000$, $\sigma = 0.1\mu$) | |
| Treatment success under BPaLC, recurrence | Gamma($\mu = 2.000$, $\sigma = 0.1\mu$) | |
| Treatment failure under BPaLM for FQ-S | Gamma($\mu = 2.000$, $\sigma = 0.1\mu$) | |
| Treatment failure under BPaLM for FQ-R | Gamma($\mu = 2.000$, $\sigma = 0.1\mu$) | |
| Treatment failure under BPaLC | Gamma($\mu = 2.000$, $\sigma = 0.1\mu$) | |

<sup>1</sup>The duration of TB symptoms when the treatment is successful was based on the time that most patients experience culture conversion (8 weeks). TB symptoms when the treatment was unsuccessful or when there was recurrence were assumed to last the full course of treatment. We assume no difference between BPaLM and BPaLC.

We also assumed that some patients with RR-TB receiving treatment experience reduced quality of life due to side effects of the treatment regimen, which we assumed to occur for the entire course of the first treatment duration. The probability of side-effects and disability weights associated with them are presented in Table 4. We used Beta distributions to model the probability and disability weights of these side-effects with variance equal to 10% the mean. Finally, we assumed that the DALYs incurred due to death are equal to the age-specific life expectancy in Moldova minus the age of the patient.

**Table 4:** Probability of adverse events of grade *≥* 3 or Severe Adverse Effects (SAE) and associated disability weights.

| Adverse event (grade $\geq 3$ or SAE) | Disability weight<sup>1</sup> | Patients with $\geq 1$ event (%)<sup>2</sup> | |
| --- | --- | --- | --- |
|  |  | BPaLM | BPaLC |
| Hepatic disorder | 0 | 12% | 5% |
| Cardiac | 0 | 2% | 4% |
| Anaemia | 0.052 | 5% | 1% |
| Creatine renal clearance decreased | 0 | 4% | 3% |
| Lipase increased/Pancreatitis | 0.114 | 3% | 2% |
| Neutropenia | 0 | 3% | 1% |
| Acute kidney injury | 0.051 | 1% | 0% |
| Lymph count decreased | 0 | 1% | 1% |

<sup>1</sup> Disability weights from supplementary appendix in [33].
<sup>2</sup> Grade $\geq 3$ or SAE by week 72 in as-treated population from supplementary appendix in [32], excluding Pneumonia and Haemoptysis.

## Results

The logistic regression model developed using the entire dataset achieved an apparent area under the receiver operating characteristic curve (AUROC) of 0.754. After correcting for optimism, the AUROC was 0.667 (95% CI [0.626, 0.717]). The calibration plot (Fig. S1, supplementary material) showed that the model underestimated the risk of resistance to FQs. Between the two calibration methods tested, Beta and Platt, the Beta calibration demonstrated better agreement between the observed and predicted risks of FQ resistance (Fig. S1), therefore predictions calibrated with Beta calibration were used in the subsequent analysis.

The current WHO guideline recommends using the BPaLM regimen for patients with TB resistant to rifampicin but without known resistance to FQs, implicitly assuming that all RR-TB cases which are not tested for FQ resistance, are susceptible to FQs. This strategy is equivalent to a risk prediction model with 0% sensitivity, correctly identifying none of the patients with FQ-resistant TB, and 100% specificity, correctly identifying all patients with FQ-susceptible TB. In terms of classification thresholds, this corresponds to using a threshold *t* = 1. Lowering the classification threshold increases sensitivity at the expense of specificity. At a threshold of *t* = 0.154, the sum of optimism-corrected sensitivity and specificity is maximized (Fig. S2, supplementary material).

To implement the PIDEMc approach, it is necessary to estimate the conditional probabilities *P* (*D*^+^ |*T* ^+^; *t*) and *P* (*D*^−^ |*T* ^−^; *t*) across a range of thresholds *t* ∈ [0, 1] (Fig. S3, supplementary material). Changing the classification threshold within this approach influences both sensitivity and specificity, which in turn affects the expected gain in NMB (panel A in Fig. 4). For a WTP value of $5714.43, equivalent to Moldova’s per capita GDP in 2022, the expected gain in NMB is maximized at a threshold of *t* = 0.308. Changing the classification threshold also affects the number of people who would be prescribed the alternative treatment regimen BPaLC (panel B in Fig. 4).

**Figure 4:**
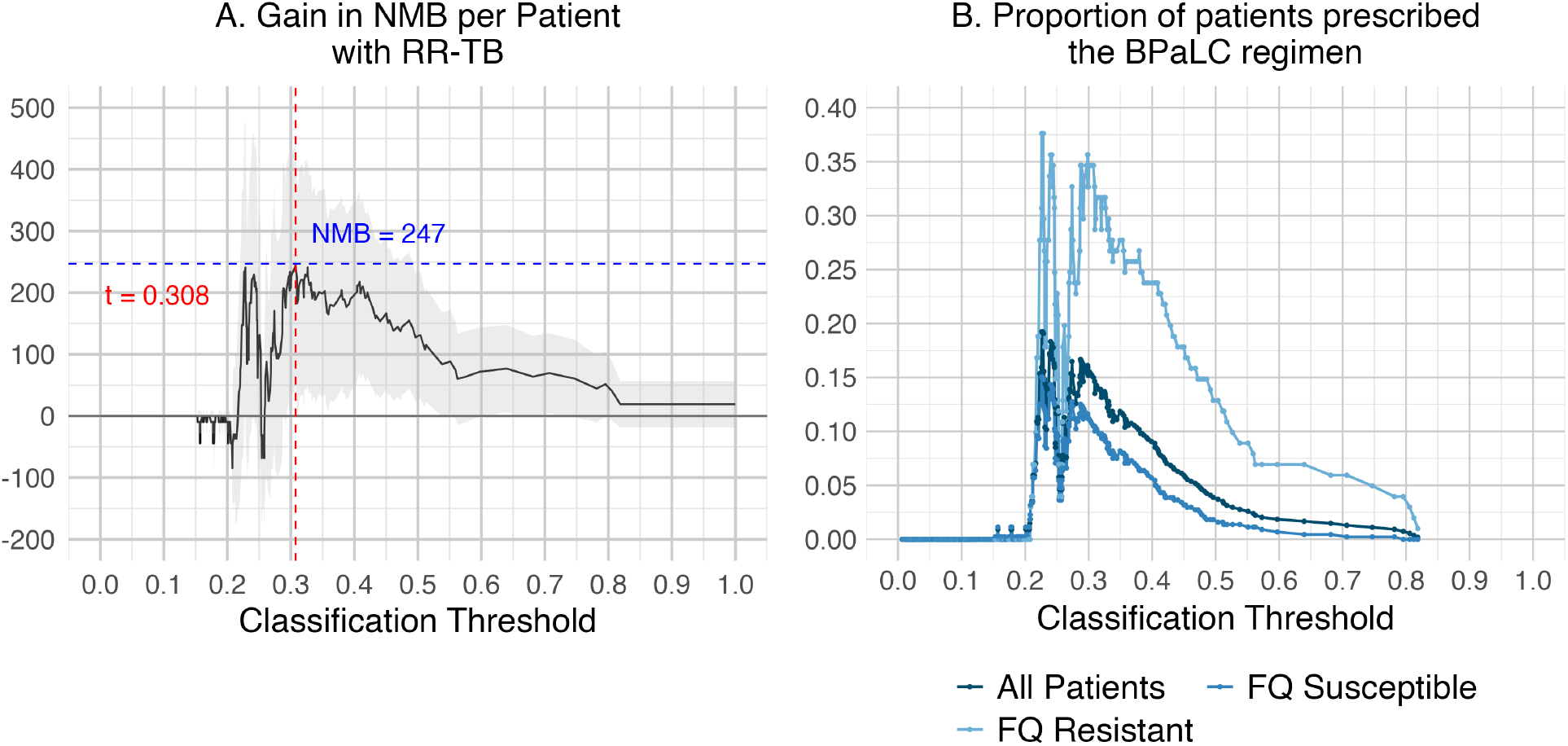
Gain in NMB by classification threshold and 95% confidence intervals (Panel A) and proportion of patients prescribed BPaLC by classification threshold and resistance status to FQs (Panel B) when PIDEMc is used with a WTP value of $5714.43, which is equivalent to Moldova’s per capita GDP in 2022.

Compared to the scenario where the WHO-recommended BPaLM regimen is prescribed for all patients with RR-TB, the personalized treatment regimens identified by PIDEMc are expected to achieve the highest gain in NMB among patients with RR-TB (panel A in Fig. 5). These gains are maintained across varying WTP values with the PIDEMc method outperforming the other methods considered. Furthermore, the PIDEMc method demonstrated robust performance even when the predictions were not calibrated; in contrast, the performance of PIDEMp and risk prediction models with arbitrary classification threshold (0.5 or a threshold that maximizes the optimism-corrected Youden’s index) deteriorated markedly when uncalibrated predictions were used (comparing panel A and B in Fig. 5).

**Figure 5:**
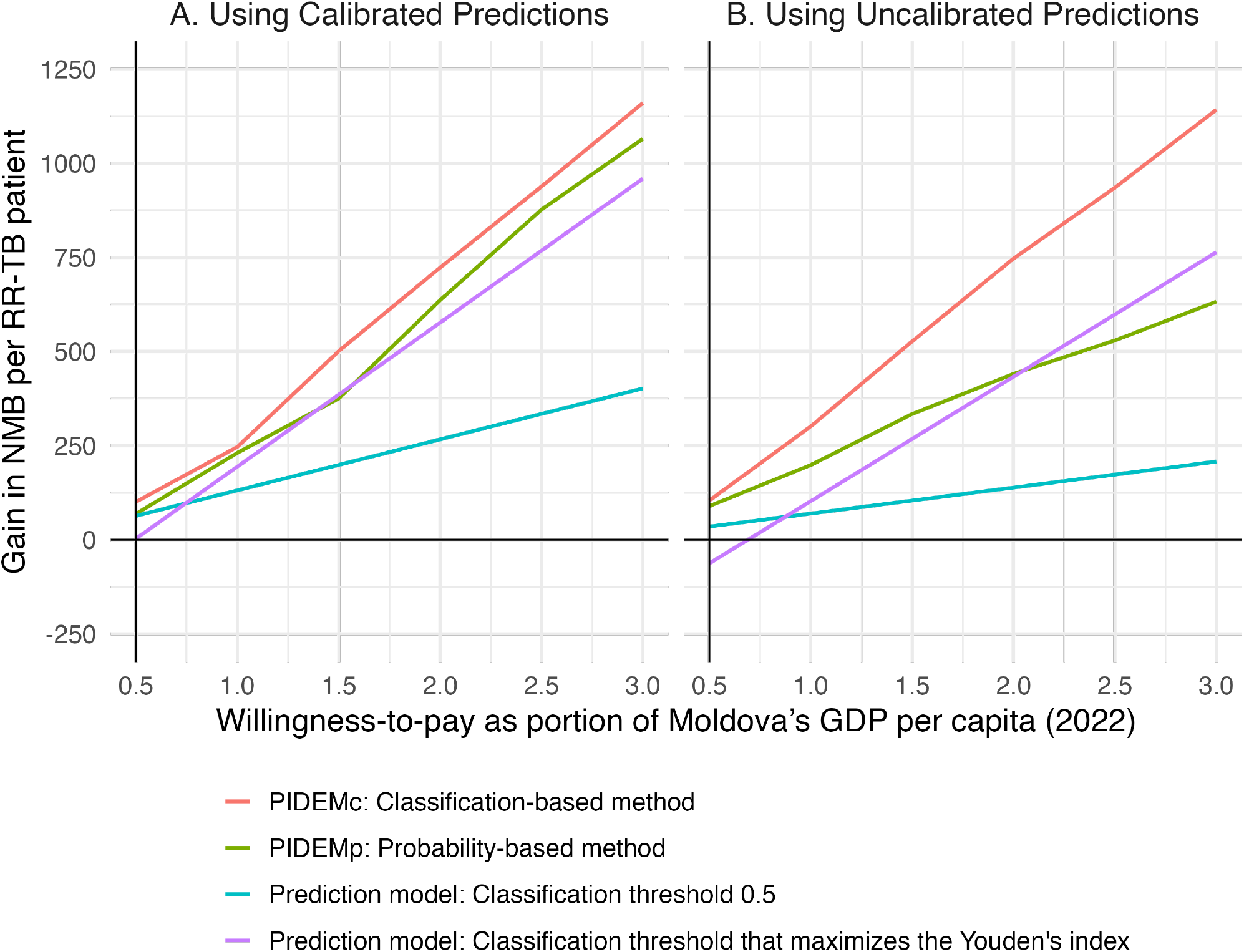
Gain in NMB for varying willingness-to-pay thresholds under different methods to inform treatment regimens compared to using the standard BPaLM regimen for all patients with RR-TB. The 95% confidence regimens along these lines are presented in Fig. S4.

Both PIDEMp and PIDEMc resulted in positive gains in NMB among patients with TB resistant to both rifampicin and FQs (Fig. 6, panel B). These gains, however, are accompanied by NMB losses among patients with RR-TB that is susceptible to FQs (Fig. 6, panel C).

**Figure 6:**
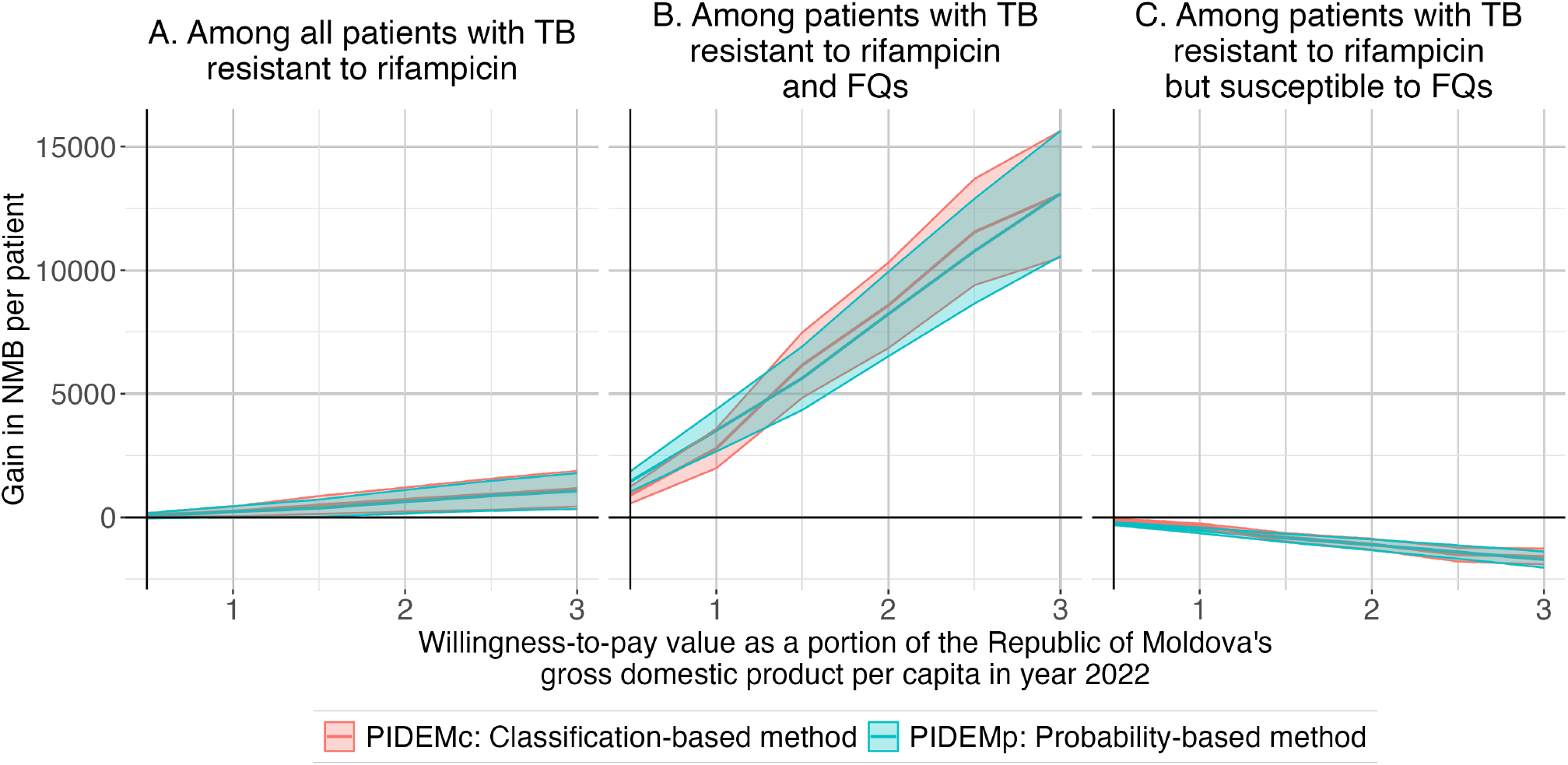
Gain in NMB among all patients with RR-TB (Panel A), among patients with RR-TB with additional resistance to FQs (Panel B), and among patients with RR-TB susceptible to FQs (Panel C) when using PIDEMc and PIDEMp to inform treatment regimens compared to using the standard BPaLM regimen for all patients with RR-TB. The shaded regions show the 95% confidence intervals.

## Discussion

Risk prediction models offer a personalized approach to support medical decision-making by using patient characteristics to predict individual-level risk. However, these models do not account for the clinical or economic consequences of their predictions (such as the costs and health impact associated with false positives or false negatives) and the recommended actions (such as the cost, efficacy, and side effects of treatment options). In this paper, we describe methods to integrate risk prediction and decision models that are designed to maximize the population NMB, accounting for the overall cost and health outcomes due to factors such as severity and duration of disease symptoms and cost, efficacy, and side effects of treatment options. We refer to these methods as Prediction-Informed Decision Models (PIDEMs). In the first method, probability-based PIDEM (PIDEMp), the risk estimate provided by a risk prediction model is used directly as a parameter in the decision model. In the second method, classification-based PIDEM (PIDEMc), the risk estimate generated by a risk prediction model is used first to classify individuals into binary disease categories, and the decision model is subsequently updated based on this classification.

We applied these methods to select between two treatment regimens for patients with RR-TB in Moldova. We found that both PIDEMp and PIDEMc resulted in higher NMB compared to the current standard of care (i.e., using the BPaLM regimen for RR-TB without known FQ resistance), and approaches based on fixed classification thresholds (e.g., a 50% cut-off or the threshold maximizing Youden’s index). Furthermore, PIDEMc resulted in a larger improvement in NMB compared to PIDEMp and its performance was not impacted markedly by whether the predictions were calibrated or not. Calibration reflects how well the predicted probabilities align with the actual risks and is a critical step in developing risk prediction models [23, 24, 25, 26]. However, perfect calibration is usually not achievable. As the PIDEMp approach incorporates predicted probabilities directly into the decision model, its performance is affected by how well the model predictions are calibrated. In contrast, the largest improvement in NMB obtained using the PIDEMc method depends on the risk prediction model discrimination, which is not affected by calibration.

A key strength of our proposed methods is the comprehensive treatment of uncertainty in both the prediction and decision modeling components. In real-world settings, the cost and effect of available medical alternatives cannot be calculated with certainty. This is mainly due to the inherent uncertainties in the decision model parameters and future factors that influence the cost and effect of the alternatives [27, 28]. For example, estimates of treatment effectiveness or the quality of life in a disease state are often derived from studies with limited sample sizes, making them susceptible to sampling error and imprecision. Likewise, estimates of risk prediction model performance metrics (e.g., sensitivity and specificity) depend on the size and quality of training data available. We used probabilistic sensitivity analysis, a standard approach in modeling studies to capture uncertainties in model input parameter [7], and bootstrap optimism correction, which is used in risk prediction modeling studies to adjust for overestimation of model performance [5], to develop a systematic approach for accounting for these sources of uncertainty.

A common approach to evaluate the utility of a risk prediction model and compare its performance with other models is decision curve analysis (DCA) [29]. DCA considers the trade-off between true positives and false positives, providing information on the net benefit of model-guided decisions. However, it does not fully account for the broader health and cost consequences associated with actions taken based on the model’s predictions. A key advantage of DCA is that it requires only the dataset used to train and evaluate the models [29]. In contrast, our proposed methods require additional data to project the health and cost consequences of using the risk prediction model to guide decision-making. Although the proposed methods require more data and computational resources, they offer a more comprehensive mechanism to evaluate and compare the utility of risk prediction models.

A limitation of our proposed methods is that we only consider scenarios with two disease states (e.g., with or without disease) and two treatment options (treat or not treat). Although this framework can be applied to many clinical decisions, extending the methods proposed to account for multiple disease states and available actions would be of immediate interest. Several risk prediction models including neural networks and random forest can predict multiple disease states and could be readily integrated into the PIDEMp approach. In contrast, extending the PIDEMc approach to accommodate multiple disease states and corresponding actions presents additional challenges, particularly in updating the probabilities within the decision model.

To maintain focus on the development and presentation of PIDEMs, we used a simple probability model to project the cost and DALYs associated with TB treatment regimens considered. Other simulation frameworks such as Markov and discrete-event simulation models are more flexible and could describe the health trajectory of patients with RR-TB more [30, 31]. Additionally, although the logistic regression model used to estimate the probability of resistance to FQs demonstrated reasonable performance, using more accurate risk prediction models with greater discriminative power is likely to enhance the gain in population NMB.

The PIDEMc method applies a single classification threshold across all patients, chosen to maximize the population NMB (Algorithm 4). In theory, however, the selection of the classification threshold could be tailored to each patient based on their preferences and other personal characteristics. Determining the classification threshold individually would require patient-level data to capture preferences over different health and cost outcomes, which is often unavailable or impractical to collect in real-world settings.

In summary, the methodologies presented in this paper provide a principled approach to identify personalized and cost-effective recommendations by integrating risk prediction and decision models. The proposed methods summarize the cost and health impact of actions informed by risk prediction models into a single measure (namely. NMB), which facilitates evaluating and comparing the utility of risk prediction models. Our proposed methods do not restrict the types of risk prediction model to estimate the probability of adverse outcomes and simulation models to project the health and cost consequence of available actions. Hence, various risk prediction and simulation models could be utilized depending on the availability of data, complexity of the disease natural history, and other practical considerations.

## Data Availability

All data produced are available online at

https://pmc.ncbi.nlm.nih.gov/articles/PMC9518704/

## Acknowledgment

Research reported in this publication was supported by the National Institute of Allergy and Infectious Diseases of the National Institutes of Health under Award Numbers R01AI177326 (to RY) and R01AI180209 (to TC). The content is solely the responsibility of the authors and does not necessarily represent the official views of the National Institutes of Health.

## A Supplementary Material

**Figure S1:**
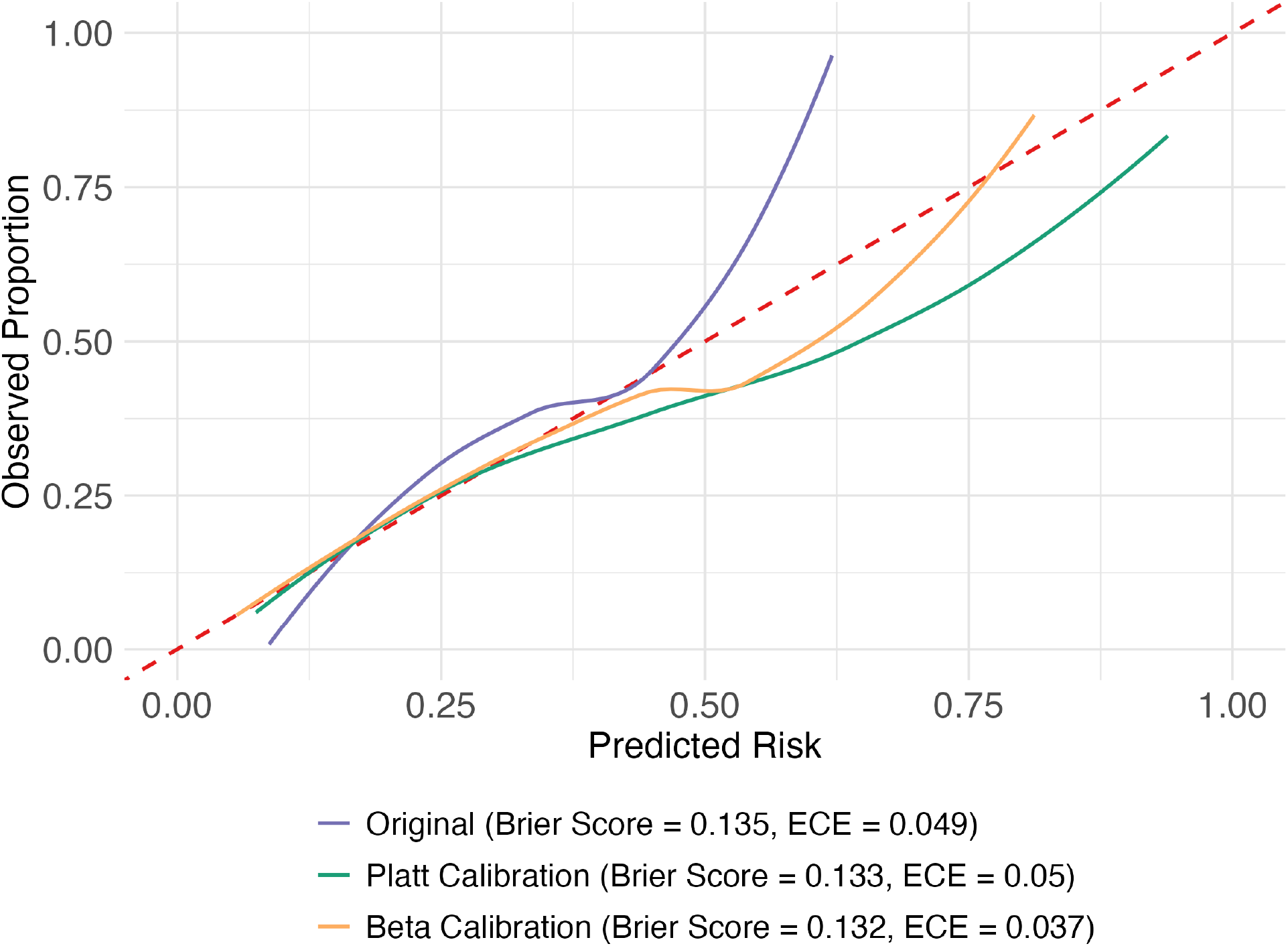
Calibration evaluation of the predictions produced by the logistic regression model using Brier score and expected calibration error (ECE).

**Figure S2:**
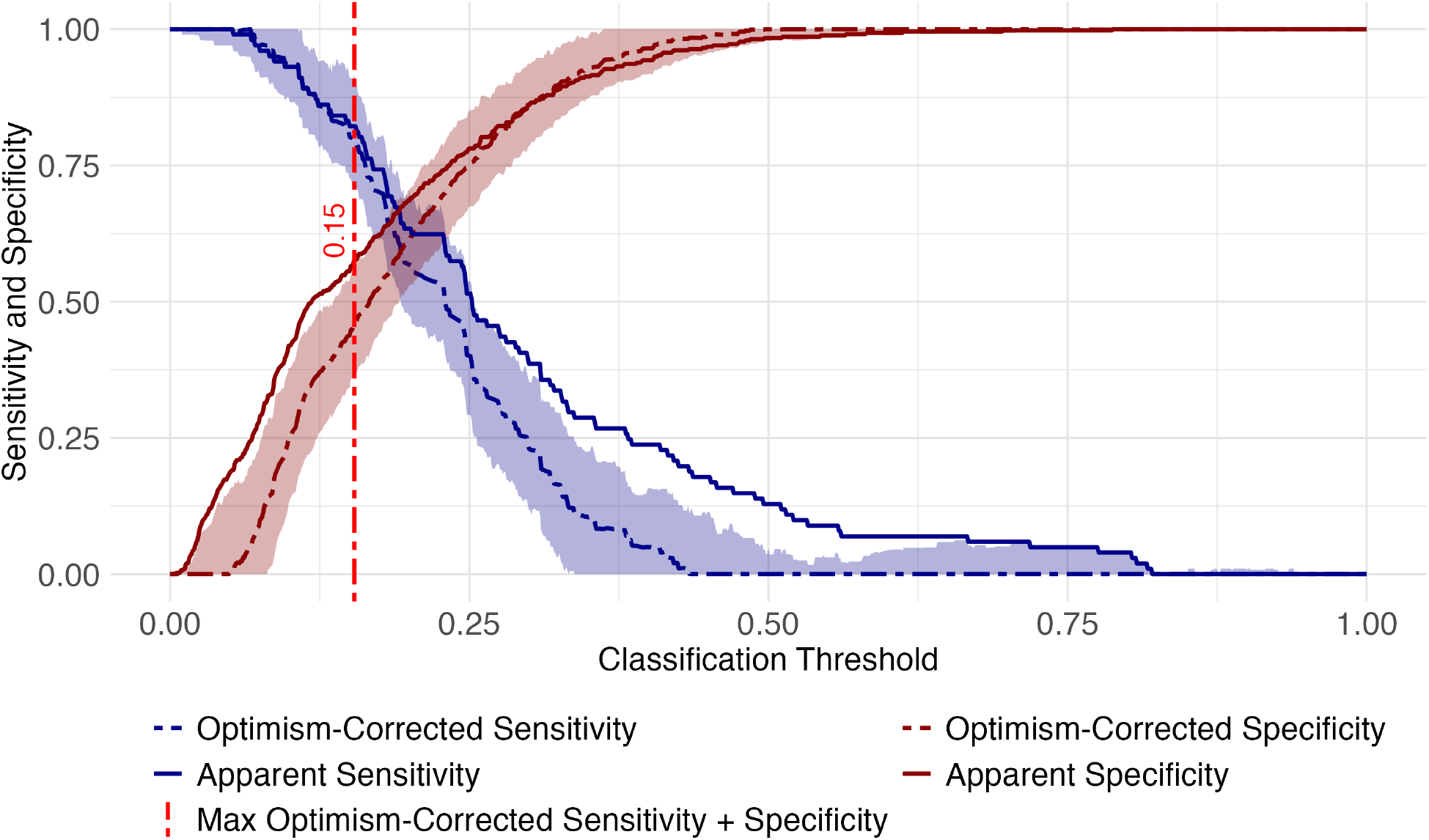
Apparent and optimism-corrected sensitivity and specificity of the logistic regression model across different classification thresholds.

**Figure S3:**
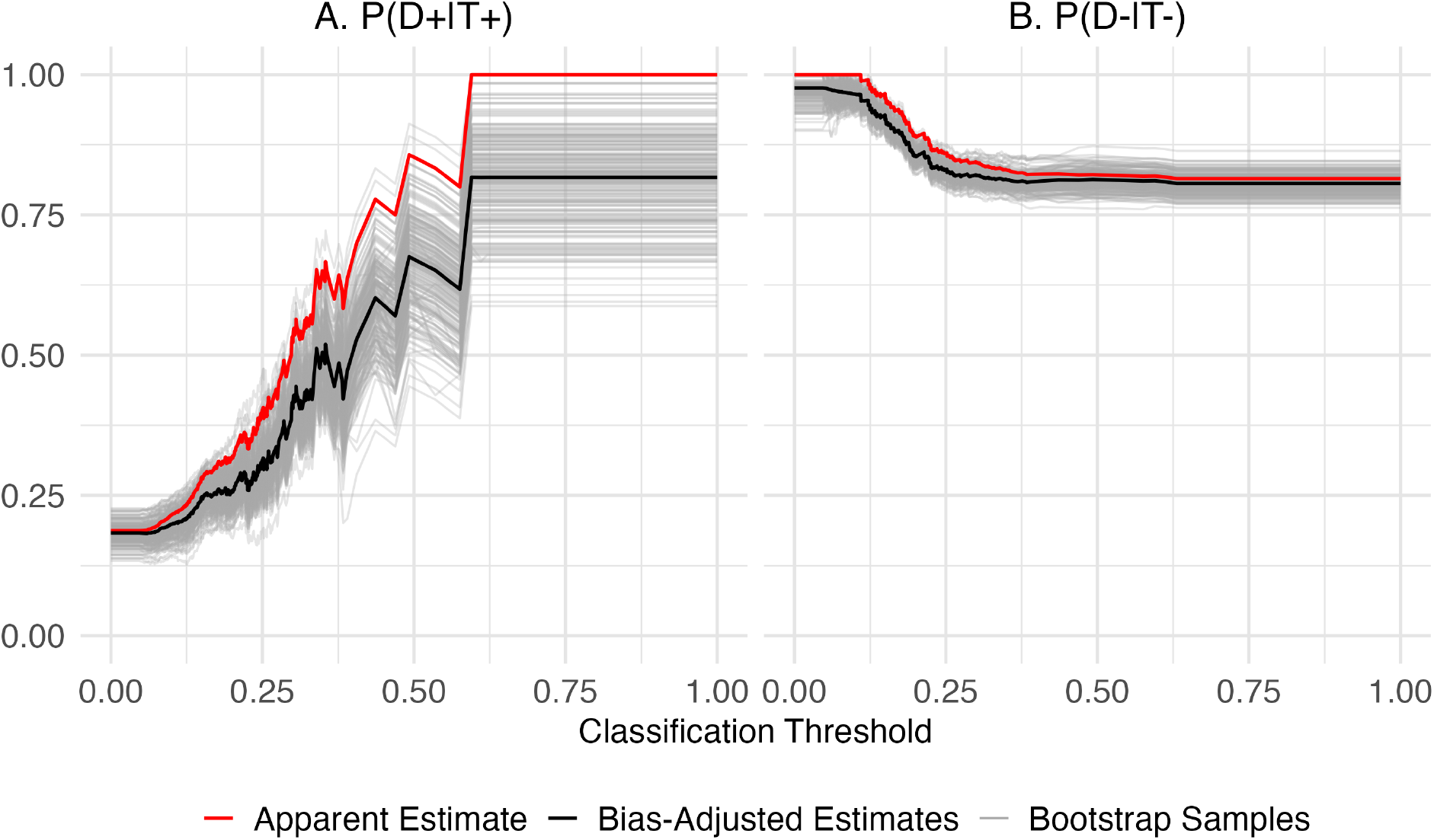
Apparent and optimism-corrected estimates for *P* (*D*^+^|*T* ^+^) and *P* (*D*^−^|*T* ^−^).

**Figure S4:**
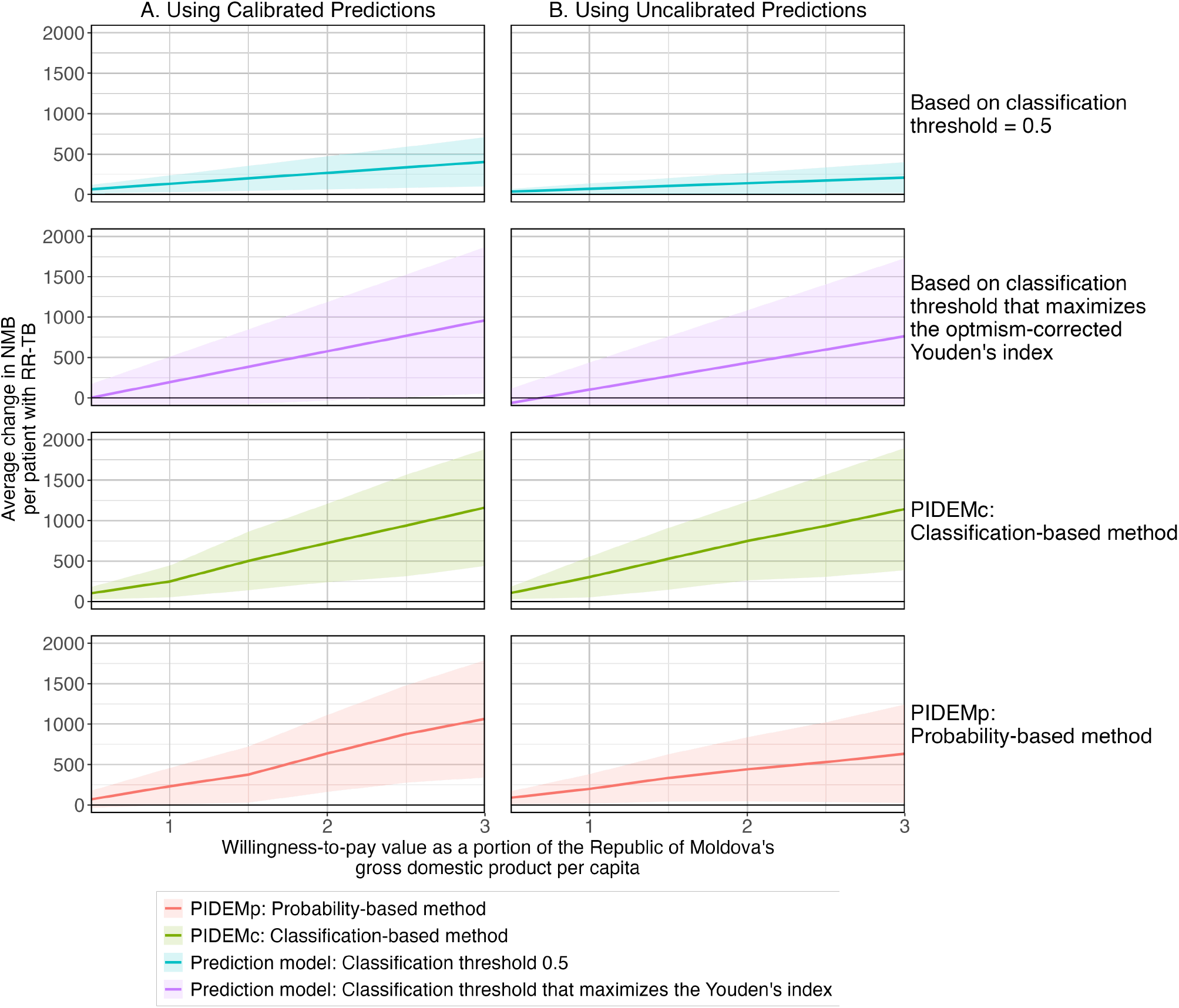
Gain in NMB for varying willingness-to-pay thresholds when proposed methods are used to inform treatment regimens compared to using the standard FQ-containing regimen for all patients with RR-TB. The shaded regions show the 95% confidence intervals.

